# Prevalence of Bicuspid Aortic Valve: Systematic Review and Meta-Analysis of Prevalence Studies

**DOI:** 10.1101/2023.06.19.23291627

**Authors:** G D. Lester, R W. Jeremy

## Abstract

Bicuspid aortic valve (BAV) is frequently reported as the most common congenital cardiac condition, with 1-2% prevalence by citing one or two autopsy studies performed up to forty years ago. Since then, echocardiography population screening studies have reported contemporaneous results, with lower prevalence, given the absence of autopsy selection bias. Therefore, we performed a systematic review and meta-analysis of autopsy and echocardiography studies to provide a more accurate synthesis of BAV prevalence. Following a systematic literature review, we performed a meta-analysis of prevalence studies using MetaXL Software Version 5.3 by EpiGear, St Lucia, Australia. The results of 45,630 autopsy patients reveal a prevalence of 0.81% (0.5-1.2%, p<0.001, n=444) and 296,245 echocardiography screened population of 0.61% (0.39-0.82% p<0.001, n=1,392) BAV prevalence. The average prevalence of 0.61-0.81% between the two studies still shows this is a common congenital cardiac condition, but shows it is less than frequently reported and thus robustly supports the case of cost effectiveness in screening first degree relatives.

## Introduction

Bicuspid aortic valve (BAV) is frequently reported as the most common congenital cardiac condition, with a 1-2% prevalence and an incidence of 1/300 (1–5). BAV is an indolent disease and often exists without the knowledge of any family history(6). However, due to the late presentation of otherwise late symptomatic disease, the actual prevalence is somewhat challenging to quantify. The recurrent citing of data showing the relatively high 1-2% unadjusted prevalence is based on the Larson & Edwards autopsy study in 1984 with 21,417 patients (7). However, by their very nature, autopsy studies have the potential for inherent selection bias when providing unadjusted data(8). Therefore, it stands to reason that population screening studies involving the echocardiography diagnosis of BAV have shown the prevalence to be considerably less, even as low as 0.1%(9).

Whilst BAV has been suggested as the most common congenital cardiac condition(10), the citation of smaller, older, and potentially biased necropsy studies in reporting prevalence, may lead to misappropriation of screening, risk and management. Furthermore, BAV exists in between 9 and 20% of a proband’s first-degree family(10,11), This led Tessler et al, to report cost effectiveness of screening first degree relatives of probands, based on a proband prevalence of 0.5-0.9%(12); however, they cite only three studies in this prevalence estimate, two of those either small (BAV n<10) or necropsy studies (13–15).

Nonetheless, the mean age of presentation of 56 years is usually the first-time symptoms of any complication occur, suggesting a disproportionate burden on a younger population relative to metabolic and degenerative aortic disease (16). Those complications include aortic stenosis, regurgitation and aneurysm(17,18). Whilst the risk of aortic complication in BAV is low at 0.2-0.5% and up to 1.2% in concomitant aortic coarctation, the prevalence of BAV results in an overall high contribution to the burden of acute aortic syndrome(19). Despite often average aortic diameters at diagnosis, between 40 and 50% of BAV patients will develop an ascending dilation, making it (assuming 1-2% prevalence) the most common cause of aortopathy (20,21). Thus, it accounts for an estimated 15% of all dissections (13,18,19). However, the outcomes in individuals with BAV are not as severe as for other genetic aortopathy, with a ten-year mortality of 3.5% (22).

To our knowledge, no meta-analysis of the prevalence of BAV has been performed. The relatively significant variation in prevalence studies of up to twenty-fold difference (of between 0.1 – 2%) depending on diagnostic methodology, may ultimately change the cost effectiveness of screening probands and their first-degree relatives; in doing so, enact monitoring and prevention to ultimately reduce morbidity and mortality. Therefore, we set out to perform a systematic review and meta-analysis of autopsy studies and compare the same research to echocardiography screening studies; this will provide a contemporary assessment of BAV prevalence to diagnose better and manage its potentially catastrophic complications.

## Methods

An international literature search was performed of studies reporting the prevalence of BAV. Google Scholar, PubMed, Scopus and ProQuest were searched in English between 1955 to Nov 2022 for the following Medical Subject Headings (MeSH) terms (23).

For the prevalence of bicuspid aortic valve with autopsy and echocardiography, the following keywords were searched:

“Aorta”, “Aorta, Thoracic”, “Dissection, Ascending Aorta”, “Dissection, Thoracic Aorta”, “Aneurysm, Ascending Aorta”, “Autopsy”, Aortic Aneurysm, Thoracic”, “Aortic Dissection”, “Bicuspid Aortic Valve Disease”, “Aortic Aneurysm, Ruptured”, “Echocardiography”, “Incidence”, “Prevalence”.

Only English language, peer-reviewed articles, were included in the review. Articles and results were reviewed by GL and RJ and excluded because they were not related to the topic or if they did not refer to any of the remaining inclusion keywords, as detailed in each PRISMA below in Figure 1.

**Figure 1.**
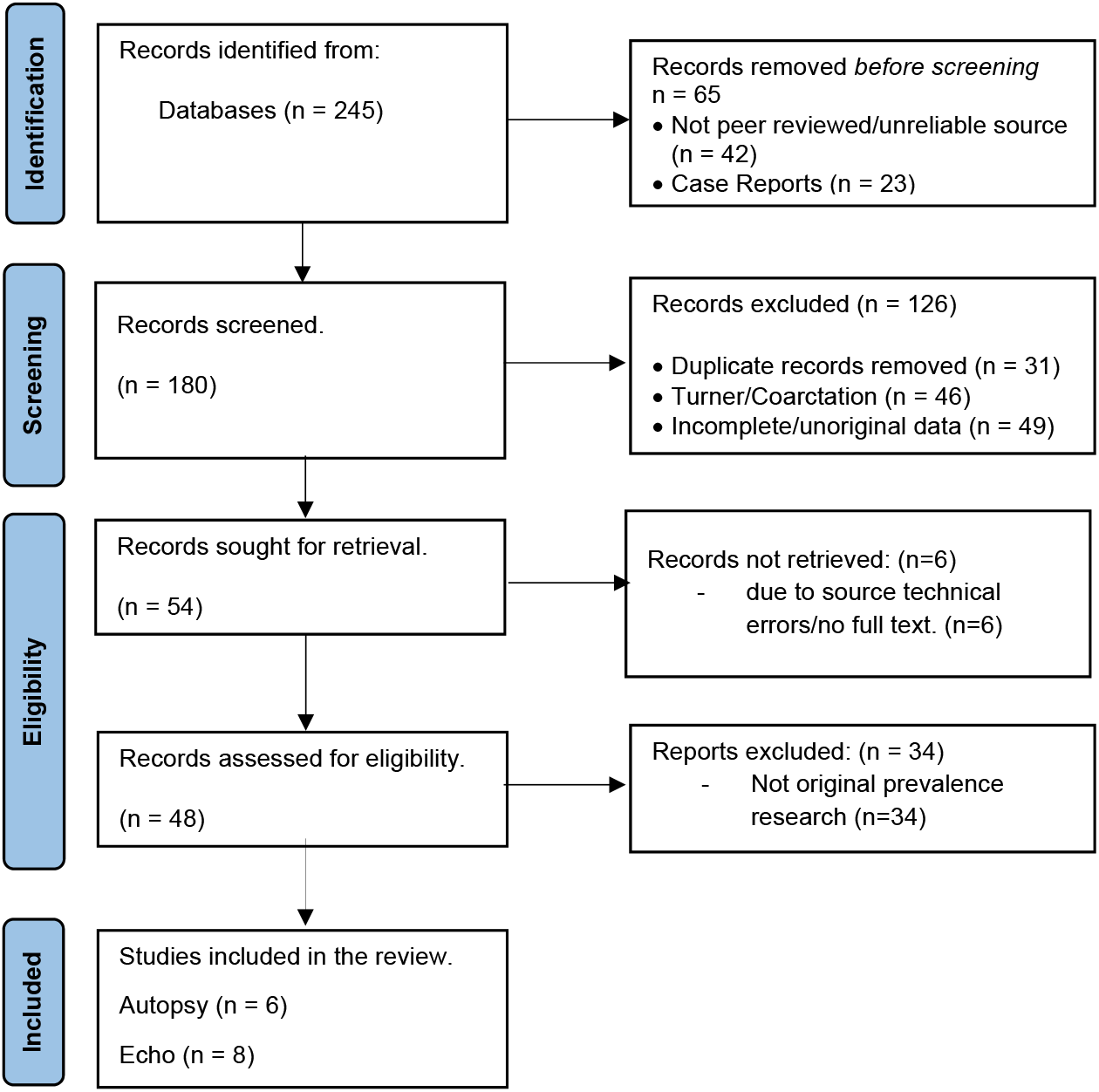
PRISMA 1: Flow Chart for systematic literature search for studies reporting the prevalence of BAV.

**Figure 2.**
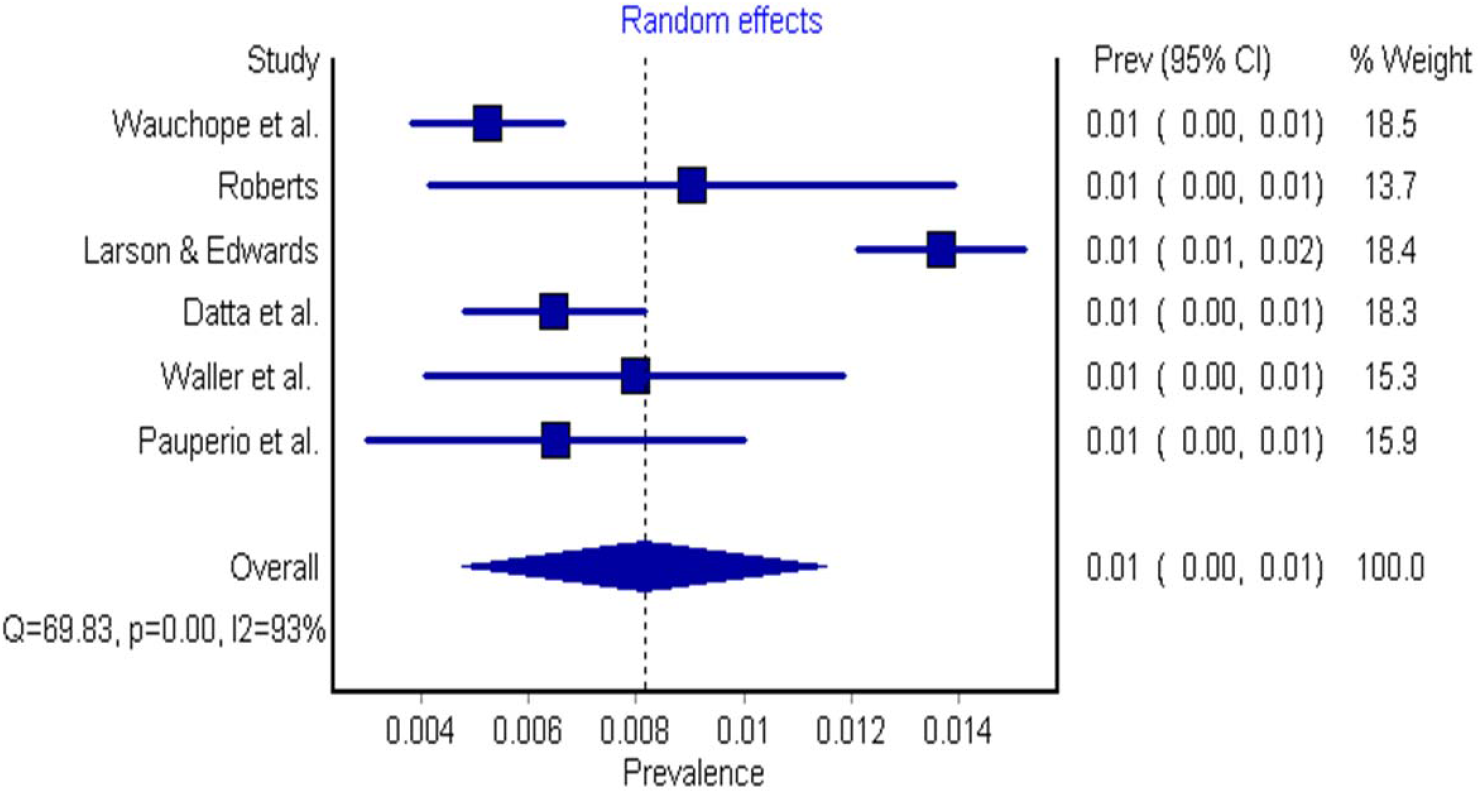
Forest Plot showing the prevalence of BAV in autopsy studies.

**Figure 3.**
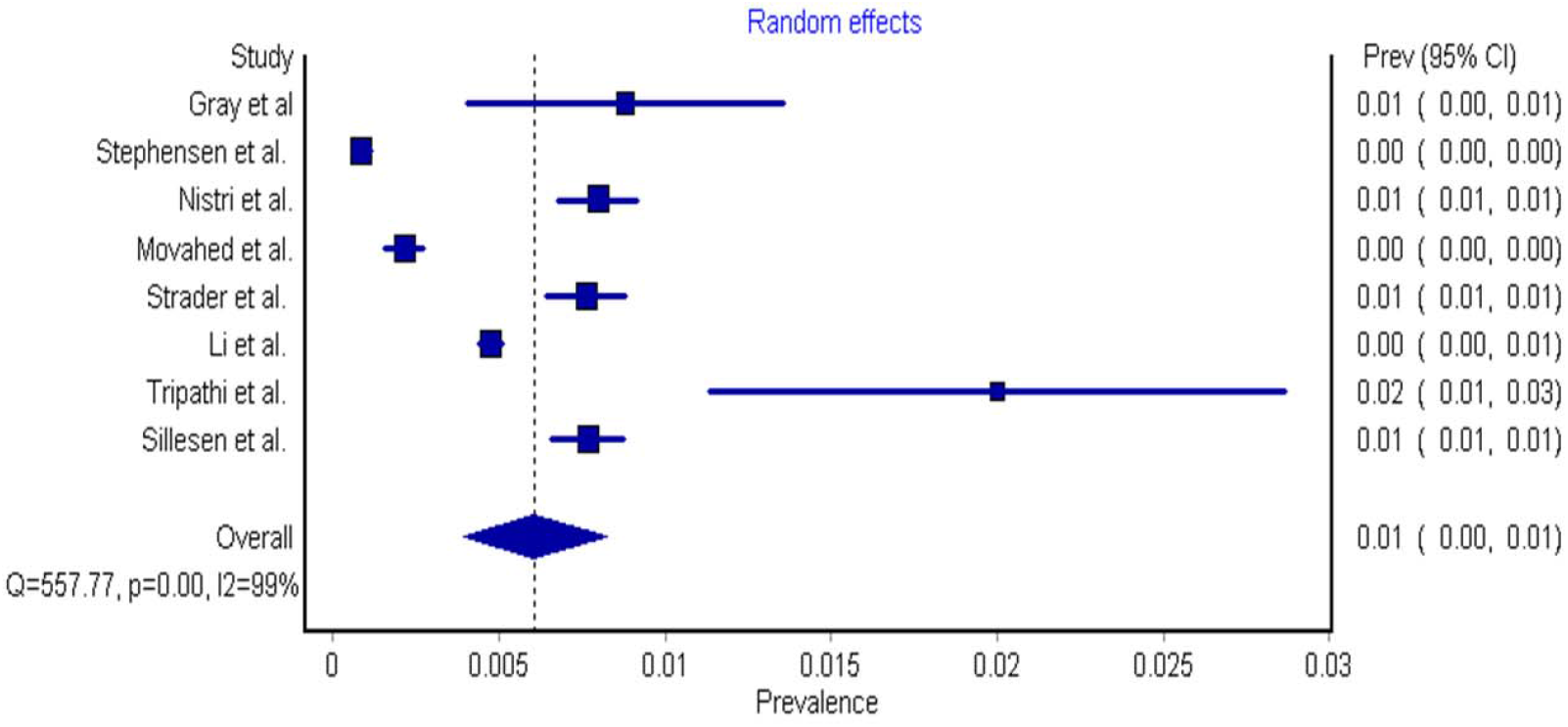
Forest Plot showing the prevalence of BAV in echocardiography studies.

The subsequent echocardiography studies identifying ten or fewer cases were excluded from the analysis, again due to the risk of sample bias: Anabwani & Bonhoeffer, Bacon & Matthews, Basso et al., Steinberger et al., Datta et al. and Tutar et al. (15,24–28).

The final fourteen articles were examined in two groups; the six autopsy studies and the eight echocardiography studies, using meta-analysis and forest prevalence plots that were performed using MetaXL Software Version 5.3 by EpiGear, St Lucia, Australia, using a random effects model in untransformed populations as described by Barendregt et al.(29–31). Statistical significance was set at 0.05%. The standard accepted definition of Bicuspid Aortic Valve was confirmed to have applied in all included cases, with two cusps clearly identified; in echocardiography studies those cusps were visible in systole and diastole in the short-axis view(32,33).

## Results

The autopsy studies are shown in Table 1 and the echocardiography studies in Table 2. There were six autopsy studies, from 1928 to 1999 from four countries; three studies from the United States, and one each from Brazil, Britain and India involving 45,630 patients with 444 diagnosed BAV cases. The largest autopsy study was the Larson & Edwards study(7), with 1.37% (n=293) diagnosed among the 21,417 patients, and the smallest was the 1970 study by Roberts of 1,400 patients, finding only 13 cases giving a prevalence of 0.9%(4). The smallest prevalence was reported by Wauchope et al. of 0.5% (n=52) of 9,966 patients(34). The mean weighted prevalence for studies diagnosed with autopsy was 0.81% (CI = 0.5-1.2%, I^2^ 92.8, p <0.001).

**Table 1.**
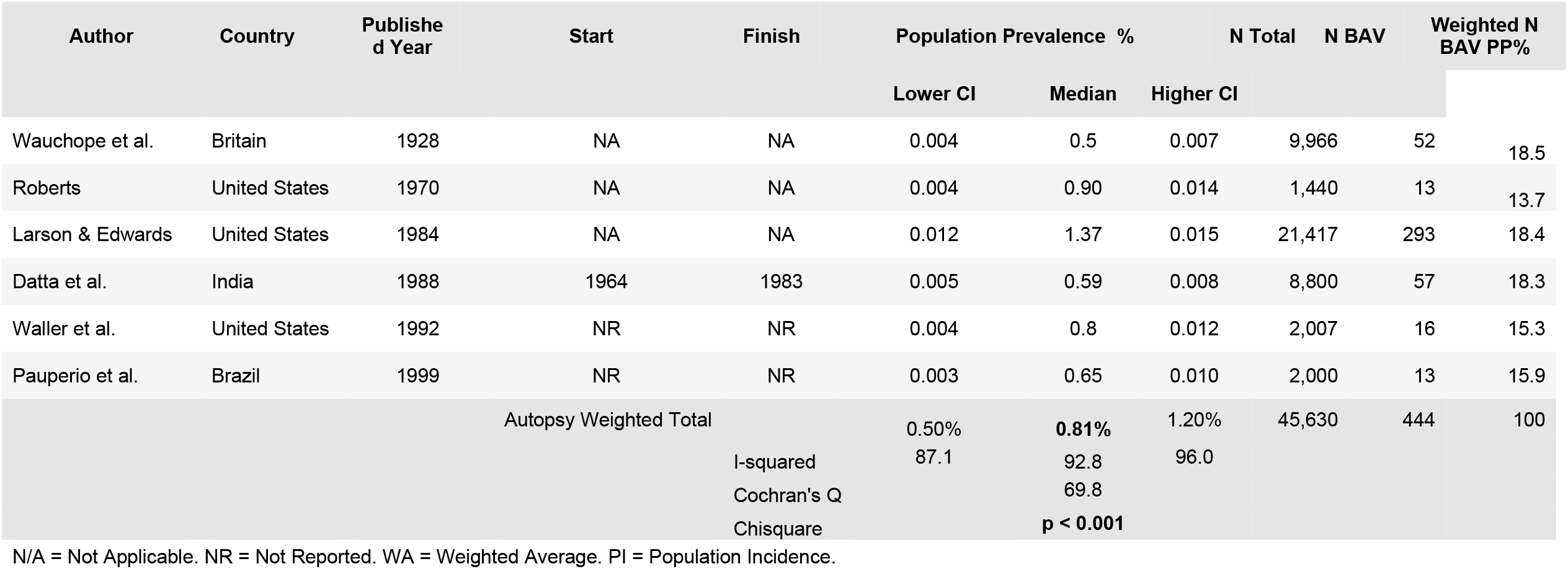
Summary of Autopsy Studies of the Prevalence of Bicuspid Aortic Valve with total calculated study weight. (4,7,26,34,44,45)

**Table 2.**
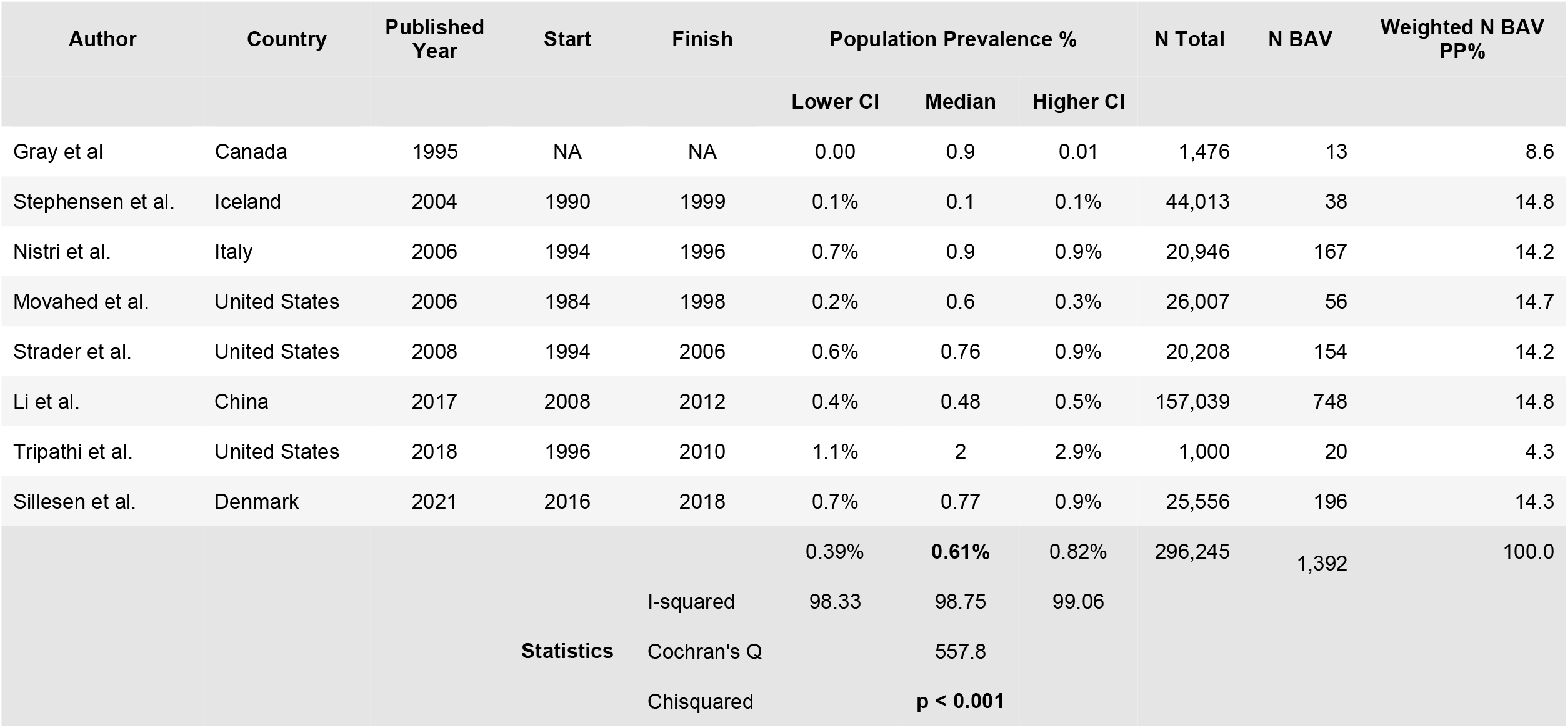
Summary of Echocardiography Sources of Prevalence of Bicuspid Aortic Valve with total calculated weighted(9,14,17,35,38,39,41,42).

There were eight echocardiography studies from six countries; three studies from the United States, and one each from Iceland, Canada, Denmark, Italy and China from the years 1995 to 2021. This involved 296,245 patients with 1,392 BAV cases diagnosed. The largest was by Li et al. in China, finding 0.48% of cases (n=748) among 157,039 people(17). The smallest study was performed by Tripathi et al., who reported 2% (n=20) of cases among 1,000 patients. With the same assumption that all individuals born with BAV survive to the time of the study, the calculated mean prevalence of echocardiography studies was 0.61% (CI = 0.39 – 0.82, I^2^ 98.7, p<0.001) (6,9,14,15,17,25,35–42).

## Discussion

To our knowledge, this is the first contemporary study to analyse all the historical studies reporting the prevalence of BAV, with combined population of 341,875 patients with 1,836 cases of BAV diagnosed. Between the two diagnostic methods examined here, there is a difference in the prevalence of BAV found in autopsy studies, of 0.81%, compared to echocardiography studies, of 0.61%, which confirms there is a higher rate of cases diagnosed by autopsy. While there is a difference of 0.2% between the two groups, the range of 0.61-0.81% is still far less than the 1-2% that is frequently reported. The significantly heterogenous I^2^ statistics (92.8 and 98.7, respectively) are consistent with the range reported in prior meta-analyses of prevalence I^2^ results (90.5-98.7) given variations in time, location, and subgroups analysed (43). However, both meta-analyses in this study revealed highly statistically significant results (p<0.001).

Whilst this remains a common congenital cardiac conditiona and thus likely the most common cause of aortopathy, the average prevalence of 0.61% - 0.81% examined in this study is less than the frequently reported figure. The cost-effectiveness of screening reported a prevalence of 0.5-0.9%(12) using three studies. Therefore, not only does this meta-analysis provide clearer guidance to clinicians and researchers about this disease frequency, it also provides more robust evidence to the cost effectiveness of screening of first-degree relatives.

## Data Availability

Full data available on request.

## Notes

### Competing Interest Statement

The authors have declared no competing interest.

### Clinical Trial

Not a clinical trial

### Funding Statement

No funding received.

